# Assessment of SARS-CoV-2 infectivity by a Rapid Antigen Detection Test

**DOI:** 10.1101/2021.03.30.21254624

**Authors:** Michael Korenkov, Nareshkumar Poopalasingam, Matthias Madler, Kanika Vanshylla, Ralf Eggeling, Maike Wirtz, Irina Fish, Felix Dewald, Lutz Gieselmann, Clara Lehmann, Gerd Fätkenheuer, Henning Gruell, Nico Pfeifer, Eva Heger, Florian Klein

**Author notes:** These authors contributed equally (Michael Korenkov, Nareshkumar Poopalasingam).

## Abstract

The identification and isolation of highly infectious SARS-CoV-2-infected individuals is an important public health strategy. Rapid antigen detection tests (RADT) are promising candidates for large-scale screenings due to timely results and feasibility for on-site testing. Nonetheless, the diagnostic performance of RADT in detecting infectious individuals is yet to be fully determined. Two combined oro- and nasopharyngeal swabs were collected from individuals at a routine SARS-CoV-2 diagnostic center. Side-by-side evaluations of RT-qPCR and RADT as well as live virus cultures of positive samples were performed to determine the sensitivity of the Standard Q COVID-19 Ag Test (SD Biosensor/Roche) in detecting SARS-CoV-2-infected individuals with cultivable virus. A total of 2,028 samples were tested and 118 virus cultures inoculated. SARS-CoV-2 infection was detected in 210 samples by RT-qPCR, representing a positive rate of 10.36%. The Standard Q COVID-19 Ag Test yielded a positive result in 92 (4.54%) samples resulting in an overall sensitivity and specificity of 42.86% and 99.89%. For adjusted Ct values <20, <25, and <30 the RADT reached sensitivities of 100%, 98.15%, and 88.64%, respectively. All 29 culture positive samples were detected by RADT. While overall sensitivity was low, Standard Q COVID-19 RADT reliably detected patients with high RNA loads. Additionally, negative RADT results fully corresponded with the lack of viral cultivability in Vero E6 cells. These results indicate that RADT can be a valuable tool for the detection of individuals that are likely to transmit SARS-CoV-2. RADT testing could therefore guide public health testing strategies to combat the COVID-19 pandemic.

**One Sentence Summary:** Standard Q COVID-19 Ag test reliably detects individuals with high RNA loads and negative results correspond with lack of viral cultivability of SARS-CoV-2 in Vero E6 cells.

## Main Text

## INTRODUCTION

Timely diagnosis of SARS-CoV-2 infection with subsequent contact tracing and rapid isolation is a critical public health strategy to contain the current COVID-19 pandemic *(1–3)*. The current gold standard of SARS-CoV-2 testing is based on real-time reverse-transcription-PCR (RT-qPCR) *(4)*. However, despite high sensitivity and specificity, RT-qPCR is less suited for rapid point-of-care identification of infectious individuals, as RT-qPCR is also able to detect non-replicating virus RNA *(5–7)*. Therefore, there is a need for an inexpensive alternative testing method to directly detect infectious individuals which can be deployed widely without the use of specialized equipment *(8, 9)*.

One promising approach is the use of lateral flow immunochromatographic assays commonly referred to as rapid antigen detection tests (RADT) designed to detect viral antigens. RADT are of particular use for community-based screenings due to low turnaround times and feasibility for on-site testing *(10, 11)*. Different tests have already been approved for clinical use, however, performance studies under real-life conditions evaluating the quality of different RADT are limited. In these studies, reported test characteristics, such as sensitivity, varied greatly depending on cohort composition (24.3-89%). While RADT showed better performance for high RNA load samples, specificity in general, however, remained high (92-100%) *(12–15)*.

As high RNA loads are typically associated with a higher probability of infectiousness, the diagnostic performance of RADT in the context of infectivity models is yet to be determined *(16– 18)*. Therefore, there is a need for large-scale field studies with a focus on virus cultivability to be able to appropriately interpret RADT results. Here, we combine side-by-side evaluation of RADT under real-life conditions in a routine diagnostic center with the cultivability of live virus from RT-qPCR positive individuals to determine the sensitivity of the Standard Q COVID-19 Ag Test (SD Biosensor/Roche) for detecting SARS-CoV-2 samples with cultivable virus in Vero E6 cells.

## RESULTS

### RT-qPCR and RADT testing in a large cohort under real-life conditions

To validate RADT performance, paired oro-and nasopharyngeal swabs were collected and tested using both RT-qPCR and RADT. RT-qPCR positive samples were additionally cultivated in Vero E6 cells to determine the ability of RADT to detect infectious samples (Fig. 1A). A total of 2,032 samples was tested with both RT-qPCR and RADT out of which 4 were excluded due to 3 missing results and 1 incorrect appliance of the RADT leaving 2,028 (99.80%) samples from 1,849 individuals eligible for analysis (Fig. 1B). SARS-CoV-2 was detected in 210 samples by RT-qPCR representing a study prevalence of 10.36%. At the time of sampling 866 (42.70%) swabs were taken from symptomatic individuals, while 810 (39.94%) specimens were collected from asymptomatic individuals. For 352 (17.35%) samples symptom status was unknown at the time of analysis. 599 (69.17%) symptomatic individuals reported up to 3 symptoms, 247 (28.52%) more than 3 and for 20 samples (2.31%) the number of symptoms was not reported. In our cohort 320 samples were obtained from 141 (7.62%) individuals who were tested twice or more (Fig. 1C). 1,239 (61.09%) samples were taken from female and 789 (38.91%) from male individuals. Participants had a median age of 32.25 years (IQR: 26.14 −43.12) (Fig. 1D). The median Ct value determined by RT-qPCR was 31.49 (IQR: 24.19 −34.16) (Fig. 1E). 126 (60%) of 210 RT-qPCR positive and 2 RT-qPCR inconclusive samples were cultivated in Vero E6 cells on the same day of sample collection. Of those 8 (4.80%) were excluded due to culture contaminations (6) or negative RT-qPCR results upon retesting (2) (Fig. 1B). A detailed cohort description can be found in the table S1.

**Fig. 1.**
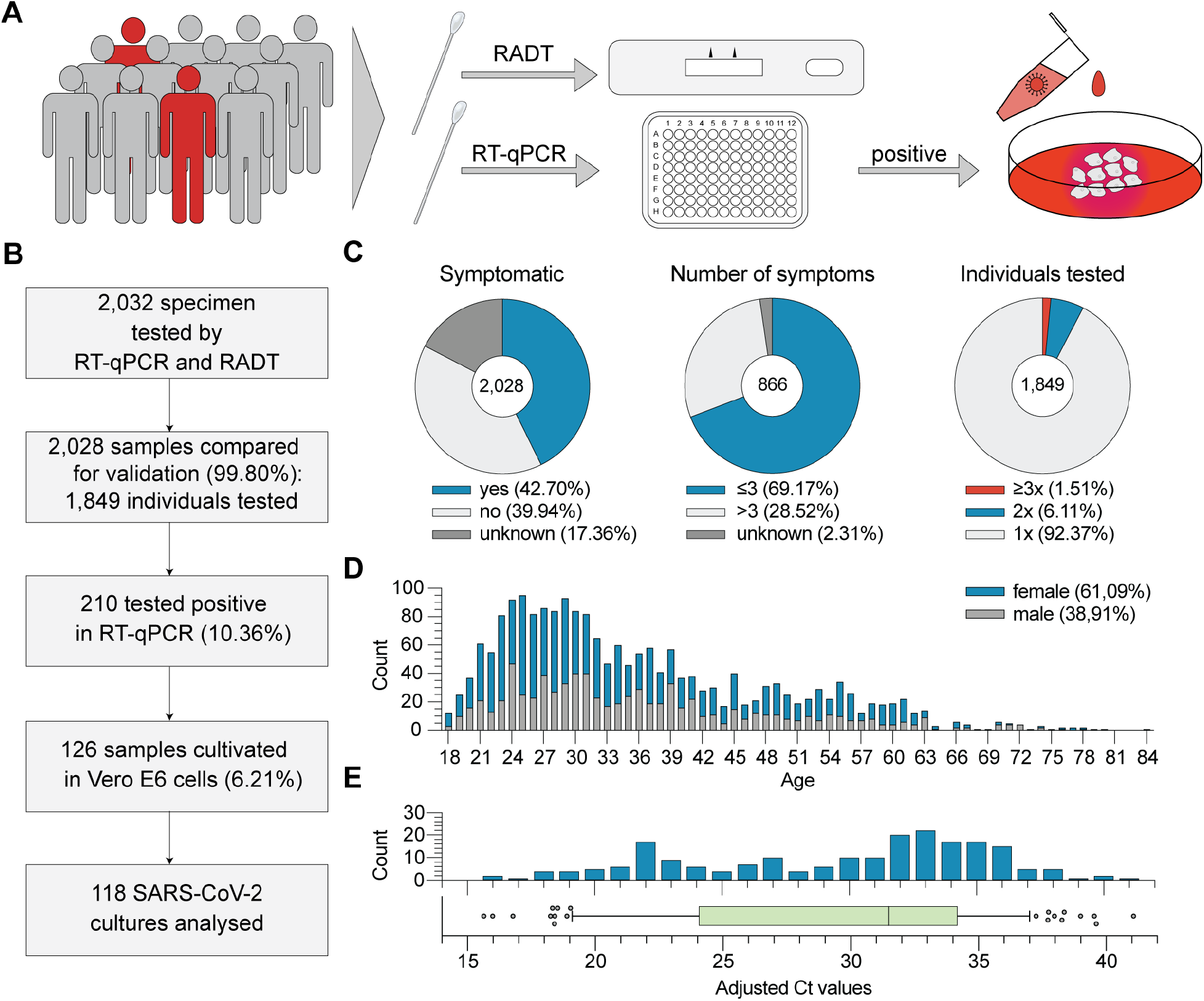
Study procedure and cohort description. **(A)** Schematic illustration of the study: Samples were obtained in parallel for both RT-qPCR and RADT testing. RT-qPCR positive samples were selected for virus culture performance. **(B)** Flowchart including cohort sizes selected for RT-qPCR, RADT and cell culture assays. Percentages in boxes 3 and 4 are referring to all analysed samples (n = 2,028). **(C)** Distribution and number of symptoms as well as test frequency among individuals. **(D)** Age and gender distribution of the cohort. **(E)** Distribution of cycle threshold (Ct) values (adjusted to cobas 6800®).

### Reliable detection of high viral load samples by RADT

The Standard Q COVID-19 Ag Test yielded a positive result in 92 (4.54%) and a negative result in 1,936 (95.46%) of all samples (Fig. 2A). Using the results of the RT-qPCR as a reference, RADT classified 90 samples (4.44%) true positive, 1,816 (89.54%) true negative, 120 (5.92%) false negative and 2 (0.10%) false positive resulting in an overall sensitivity and specificity of 42.86% and 99.89%, respectively (Fig. 2B). For positive RADT results the median Ct was 23.32 (IQR: 21.48 −26.69) with a median copy number/ml of 6.69 log_10_ (IQR: 5.57 log_10_ – 7.3 log_10_) compared to 33.46 (IQR: 32.04 −35.38) and 3.3 log_10_ (IQR: 2.66 log_10_ – 3.79 log_10_) for negative RADT results (p<0.0001; Fig. 2C). Stratified by adjusted Ct values the RADT had sensitivities of 100% (14/14), 98.25% (56/57), 88.64% (78/88) and 50.57% (89/176) for adjusted Ct values <20, <25, <30, <35, respectively (Table 1). Diagnostic sensitivities of 93.33%, 55.55% and 22.22% are reached for adjusted Ct values between 25-26, 27-28 and 29-30 and sensitivities of 86.36%, 29,17% and 9.68% for 6 log_10_, 5 log_10_ and 4 log_10_ copies/ml, respectively (Fig. 2D). We conclude that observed RADT sensitivity declines at adjusted Ct values above 27 or below 6 log_10_ copies/ml but reliably detects samples with higher RNA loads.

**Table 1.**
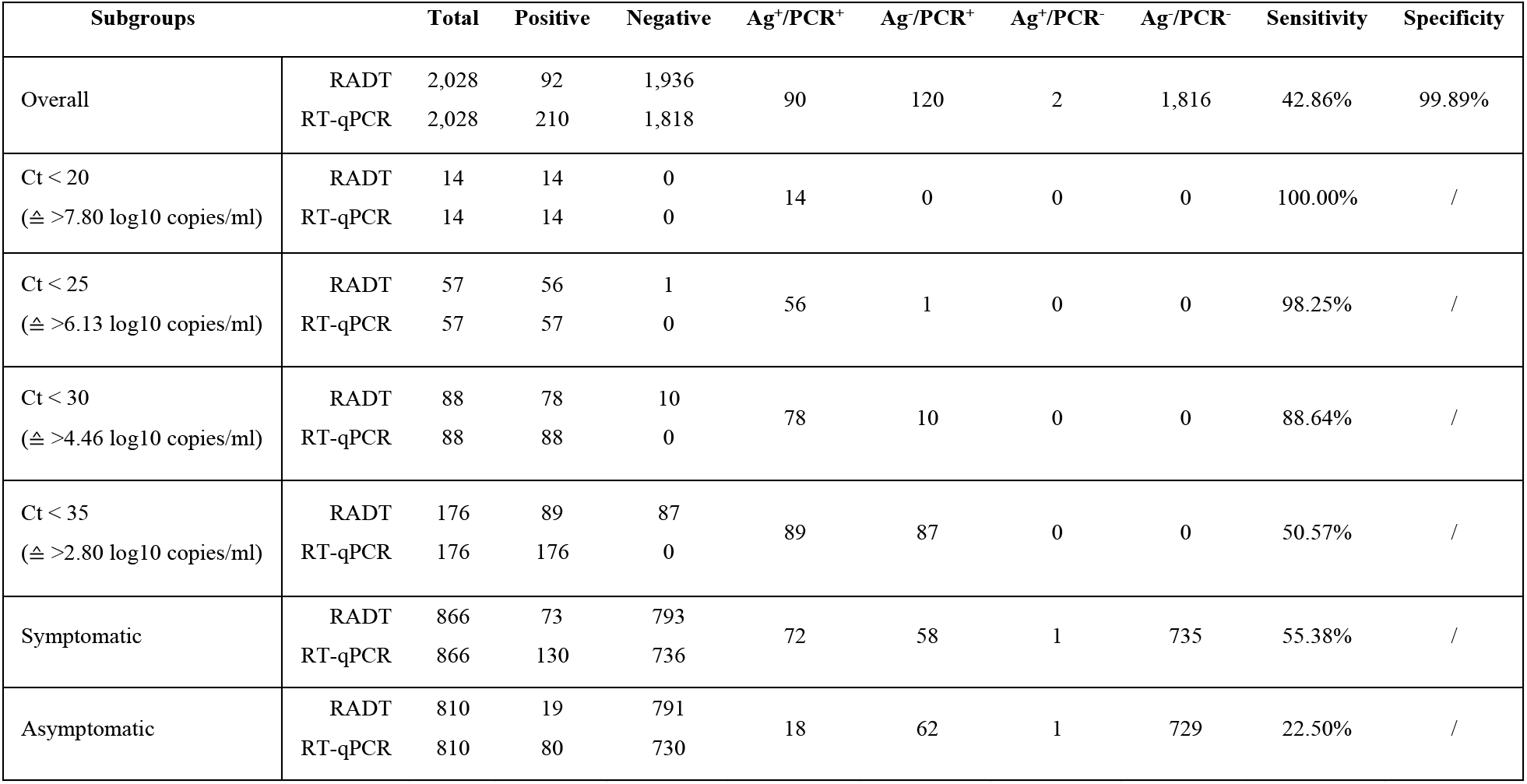
Performance data of the Standard Q COVID-19 Ag Test. Test results and head-to-head comparison of RADT and RT-qPCR overall and for different subgroups.

**Fig. 2.**
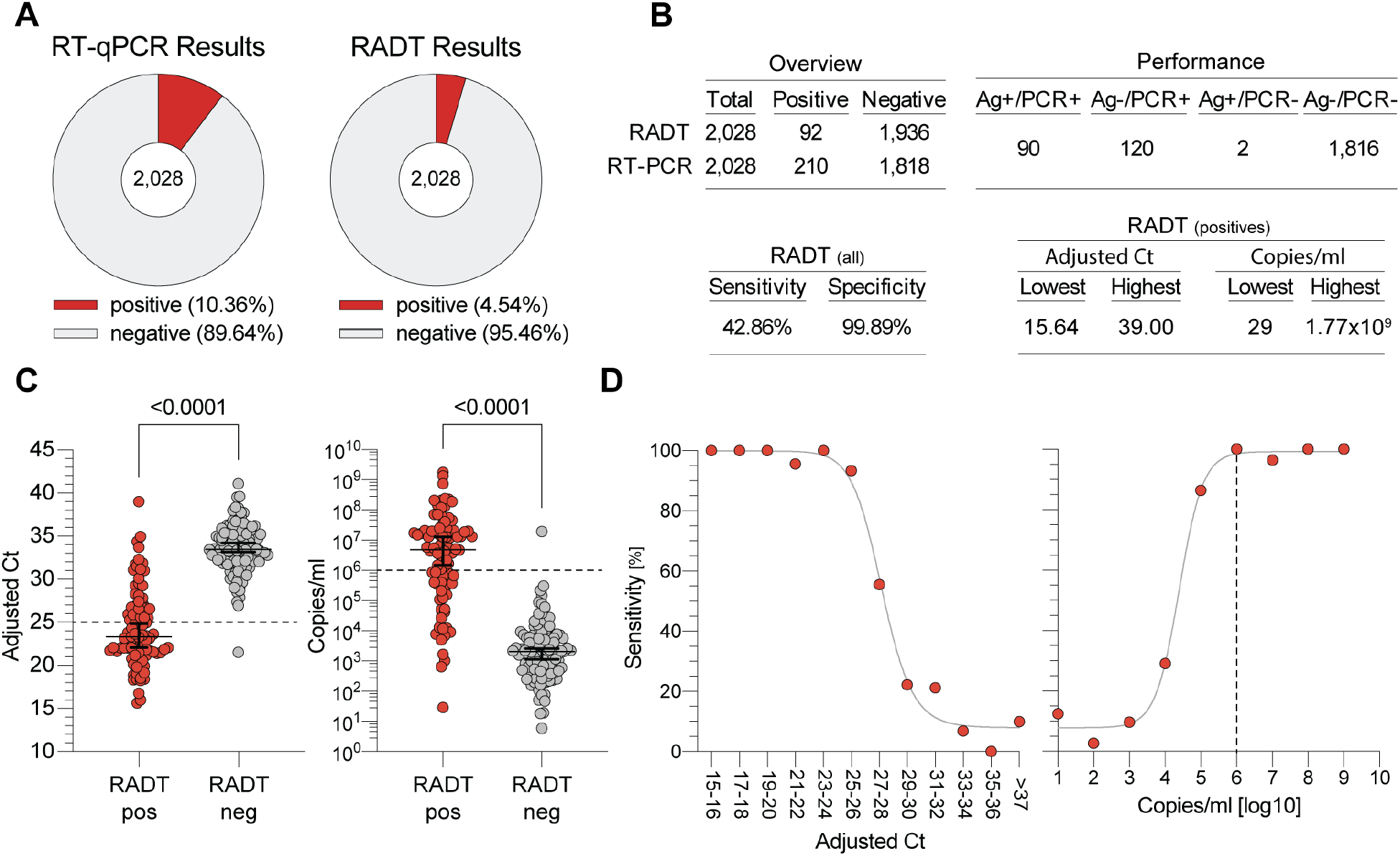
Head-to-head comparison of SARS-CoV-2 detection by RT-qPCR and RADT. **(A)** RT-qPCR and RADT results of all 2,028 specimens. **(B)** Performance data of the Standard Q COVID-19 Ag Test. **(C)** The 210 RT-qPCR positive samples are plotted by adjusted Ct values and SARS-CoV-2 RNA load, respectively, and stratified by their RADT result (p < 0.0001, MWU). **(D)** The sensitivity of the RADT is stratified by adjusted Ct values and RNA load in log copies/ml. The graph on the left shows data points for two Ct units each. Due to low sample sizes, values > Ct 37 were combined.

### Decreased RADT sensitivity over the course of symptom duration

Data on symptoms was obtained and analyzed for 1,676 (82.64%) out of 2,028 samples. 130 (15.01%) and 73 (8.43%) of 866 samples from symptomatic subjects were tested positive in RT-qPCR and RADT, respectively (Fig. 3A). Symptom duration at the time of sampling was reported for 860 (99.31%) samples with a median duration of 2 days since symptom onset for both RT-qPCR positive (IQR: 1 −6) and negative (IQR: 1 −4) samples (Fig. 3B). Of samples tested positive by RT-qPCR, RADT detected 56 (68.29%) samples within 4 days since symptom onset. When reported, median symptom duration for RT-qPCR positive samples with either a RADT positive (n=69) or negative (n=55) result was 2 days (IQR: 1 −3; Fig. 3C). For samples from symptomatic patients the median adjusted Ct and median copies/ml were 28.90 (IQR: 23.17 −32.80) and 4.83 log_10_ (IQR: 3.53 log_10_ – 6.74 log_10_), respectively. In individuals not reporting any symptoms, detected viral load was significantly lower (median adjusted Ct: 33.45 (IQR: 31.26 −35.31); median copies/ml: 3.31 log_10_ (IQR: 2.69 log_10_ – 4.05 log_10;_ p<0.0001; Fig. 3D). Symptoms were reported in 3/6 (33.3%), 30/62 (48.39%), 21/22 (95.45%) and 29/39 (74.36%) cases for samples under 2 log_10_, 3-4 log_10_, 5-6 log_10_ and over 7 log_10_ copies/ml, respectively (Fig. 3E). Samples with RNA concentrations above 6 log_10_ were only observed up to 6 days after symptom onset. 41 of 42 samples above 6 log_10_ (97.62%) were tested positive by RADT. After 8 days since symptom onset 2 of 22 specimens were detected by RADT, resulting in a sensitivity of 9.09%. Thus, we conclude that the sensitivity of the RADT decreased with symptom duration as viral loads declined (Fig. 3F, G).

**Fig. 3.**
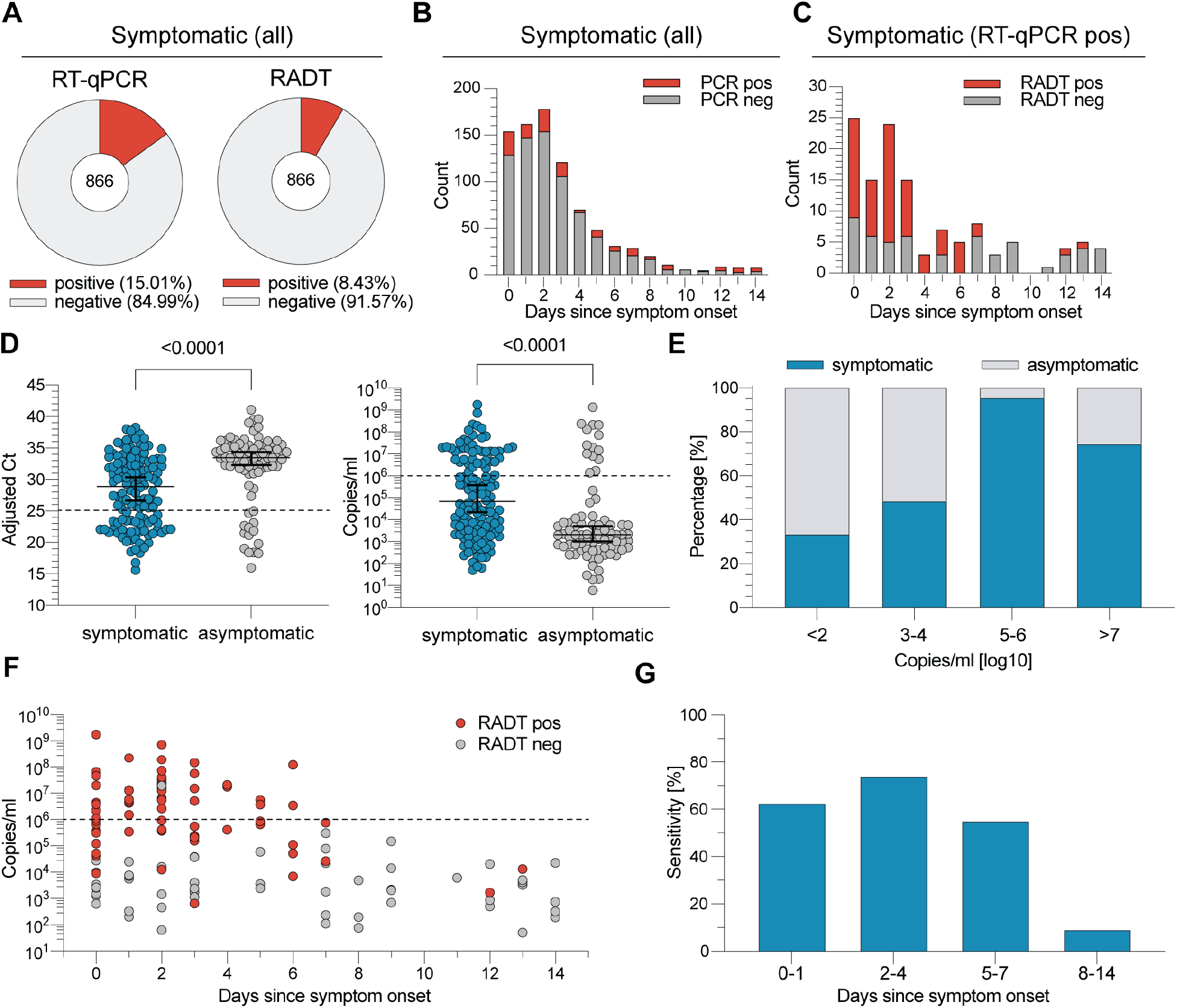
RT-qPCR and RADT results in symptomatic and asymptomatic individuals. **(A)** RT-qPCR and RADT results of the 866 samples from symptomatic participants. **(B)** The number of RT-qPCR positive and negative specimens from symptomatic individuals is plotted by the days since symptom onset. **(C)** The number of RADT positive and negative specimens within the RT-qPCR positive samples of symptomatic participants (n = 130) is stratified by the days since symptom onset. **(D)** Symptomatic and asymptomatic RT-qPCR positives are plotted by adjusted Ct value and RNA load (p < 0.0001, MWU). **(E)** The Proportion of samples from symptomatic and asymptomatic individuals is stratified by SARS-CoV-2 RNA loads. **(F)** RT-qPCR positive samples of symptomatic individuals are plotted by RNA load and stratified by the number of days since symptom onset. The RADT result is indicated in corresponding colors. **(G)** RADT sensitivity is stratified by days since symptom onset.

### Negative RADT result corresponds with lack of viral cultivability

From 118 inoculated cultures CPE was observed in 29 (24.58%) (Fig. 4A, B). To confirm virus replication RT-qPCR of culture supernatant taken on day 4 and 7 was performed as described previously and compared to the initial RT-qPCR result from swab medium. Observed CPE and RT-qPCR results matched in 116 (98.30%) cases and mismatched in 2 cases (Fig. 4C). The RT-qPCR results were used for further analysis leaving 29 (24.58%) SARS-CoV-2 positive cultures. Initial Ct values and copies/ml of positive cultures ranged from 15.64 to 24.97 and 6.14 log_10_ to 9.25 log_10,_ whereas negative cultures ranged from 21.5 to 38.27 or 1.71 log_10_ – 7.30 log_10,_ respectively (Fig. 4D). All (29/29) positive SARS-CoV-2 cultures had been previously identified as positive by RADT. Of 89 negative cultures 64 (71.91%) had been previously classified as RADT negative and 25 (28.09%) as RADT positive resulting in a sensitivity of 100.00% and a specificity of 71.91% for detecting viral cultivability by RADT (p<0.0001; Fig. 4E). Therefore, the calculated positive predictive value (PPV) of RADT for viral cultivability in Vero E6 cells was 54.72% and the negative predictive value (NPV) 100% (Table 2). For RADT and culture positive results the median adjusted Ct was 20.8 and median RNA load in copies/ml was 7.53 log_10_ compared to 33.39 and 3.34 log_10_ for RADT and culture negative results. Median adjusted Ct and copies/ml for RADT positive but culture negative results were 25.43 (IQR: 23.39 −27.72) and 5.99 log_10_ (IQR: 5.23 log_10_ – 6.67 log_10_), respectively (Fig. 4F). With viral RNA declining over the course of disease the ability to isolate virus decreased with no positive culture after 6 days since symptom onset (Fig. 4G). To confirm our SARS-CoV-2 culture results, 6 dilution series with sample Ct values ranging from 15.5 to 17.64 were prepared. Two series were excluded due to no recovered virus in one case and failed RT-qPCR of culture supernatant in the other. Virus cultivation and RADT testing from 4 remaining dilution series showed that all SARS-CoV-2 positive cultures were previously detected by RADT (fig. S1). Probit regression of RADT and cell culture analyses show greater probabilities of positive result (PPR) for low adjusted Ct values and high RNA loads, respectively. The virus culture assay shows 90% and 50% PPR for an adjusted Ct value around 21.45 or 7.31 log_10_ copies/ml and 23 or 6.8 log_10_ copies/ml, respectively. RADT show a PPR of 90% and 50% at an adjusted Ct value of 24.7 or 6.24 log_10_ copies/ml and 29.0 or 4.78 log_10_ copies/ml, respectively (Fig. 4h). In summary these data show that a negative RADT result can reliably predict non-infectiousness in our culture Vero E6 assay.

**Table 2.**
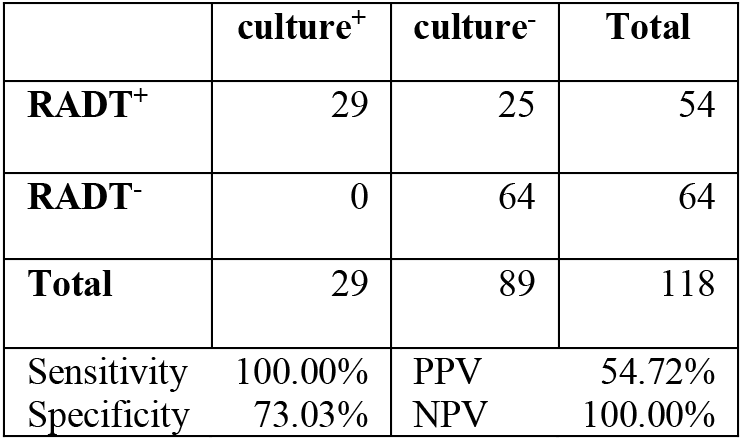
Comparison of RADT and culture results. Analysis of RADT performance in the context of culture infectivity (p < 0.0001, Fisher’s exact test; weighted analysis p < 0.0001).

|  | culture<sup>+</sup> | culture<sup>-</sup> | Total |
| --- | --- | --- | --- |
| **RADT<sup>+</sup>** | 29 | 25 | 54 |
| **RADT<sup>-</sup>** | 0 | 64 | 64 |
| **Total** | 29 | 89 | 118 |
| Sensitivity | 100.00% | PPV | 54.72% |
| Specificity | 73.03% | NPV | 100.00% |

**Fig. 4.**
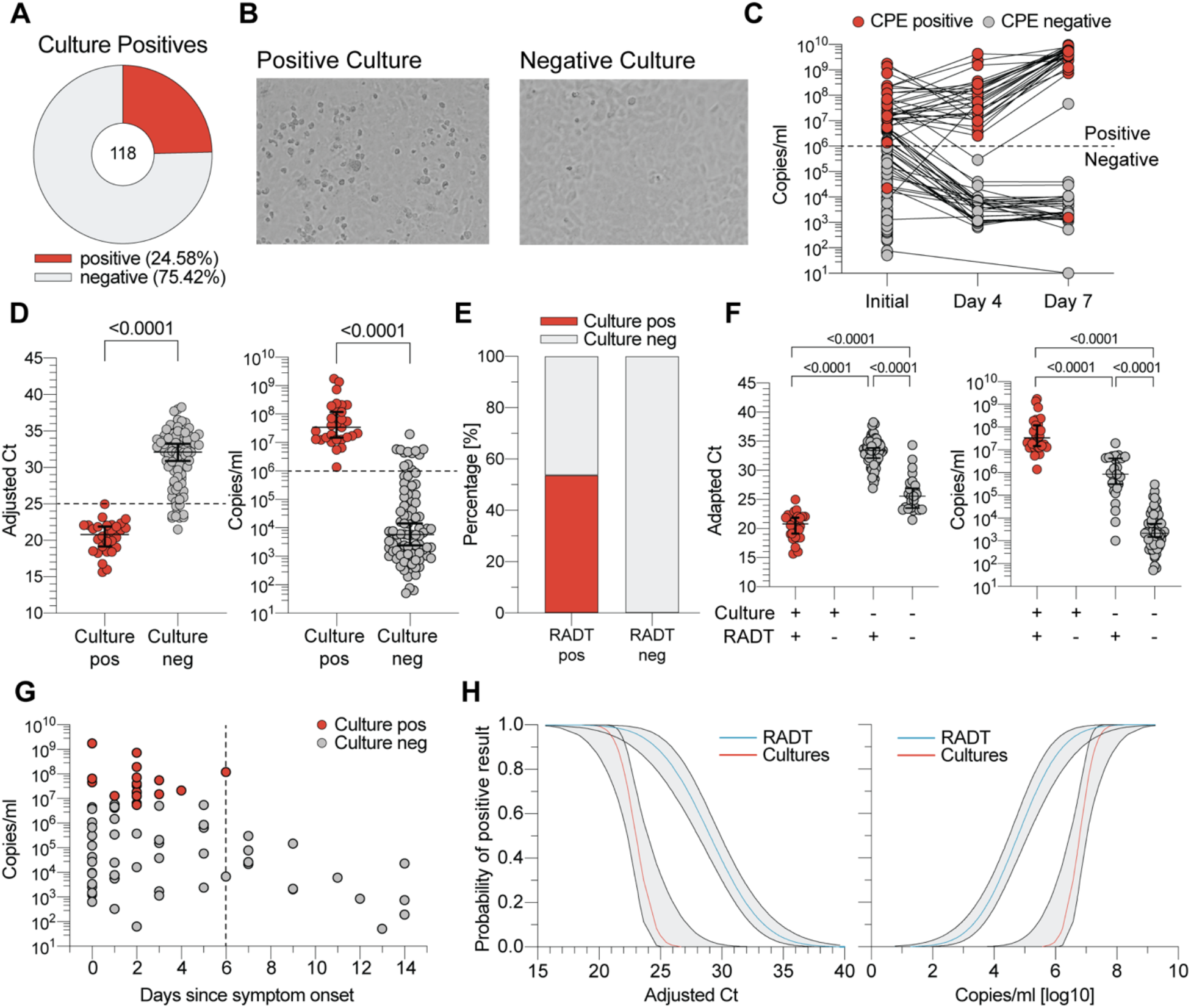
Virus culture analysis and RADT performance in the detection of replication competent SARS-CoV-2. (**A**) Proportion of culture positive specimens. **(B)** Exemplary images of cultures positive and negative for cytopathogenic effect 7 days post inoculation. **(C)** RNA load comparison of swab medium (= initial) and virus culture supernatant on days 4 and 7 to determine virus replication by RT-qPCR. CPE result is indicated in corresponding colors. **(D)** Positive and negative viral cultures plotted by adjusted Ct and RNA load of the original swab medium (p < 0.0001, MWU). **(E)** Proportion of positive and negative virus cultures by RADT result. **(F)** Adapted Ct values and RNA loads of cultured samples stratified by culture and RADT result (p < 0.0001, MWU). **(G)** Culture positive and negative samples are plotted by RNA load of original swab medium and stratified by the days since symptom onset. **(H)** Probability of positive result for RADT and viral cultures in the context of adjusted Ct values and RNA load (Probit-Model, R-function GLM).

### Adjustments for repeated testing reveal no statistically significant difference

To evaluate whether repeated testing of several individuals affects our conclusions, additional statistical analyses were carried out (see Material and Methods). The results shown in Fig. S2 confirm conclusions from Fig. 2B, 2D and 4H, respectively. P-values for Table 2 and Table S1 and S2 were calculated with Fisher’s exact test. Correction for repeated RADT measurements was based on weighted counts (cf. Fig. S2A) rounded to whole numbers, yielding very similar p-values to the unweighted variant (Fig. S1 and S2; in brackets). In addition, the performance of the RADT was analyzed for the first measurement of each subject only (simplified analysis). Calculated sensitivities showed almost identical values as overall sensitivities described above (Fig. S2C). For all studies that applied an MWU-test (Fig. 2C, 3D, 4D, 4F), the p-values remain p<0.0001 according to GEE.

## DISCUSSION

RADT are cheap and fast diagnostic tools that can be immediately performed at the point of care. Here, we present comprehensive data on the use of Standard Q COVID-19 RADT for high throughput testing of a large cohort tested under real-life conditions in a SARS-CoV-2 outpatient diagnostic center.

Implementation of the RADT on top of routine diagnostic was completed without much difficulty as the rapid feasibility made it easy to run multiple tests in parallel for a single operator. Upon following the manufacturer’s instructions there were only two cases in which the result could not be read out clearly. As a consequence, we were able to conduct 2,028 paired RT-qPCR and RADT tests directly on site and cultivate virus on the same day without prior sample freezing. While sensitivities of other RADT vary between 24% and 93% in different studies *(12, 19–21)*, reported Standard Q RADT sensitivities are mostly in the range from 68% to 90% *(13, 22, 23)*. The overall diagnostic sensitivity observed in our study was 42.86%. However, the investigated cohort of non-hospitalized patients was to a large extend comprised of individuals with adjusted Ct values over 30 (122/210) who were still detected in RT-qPCR due to persisting viral RNA. Stratification by RNA load revealed that the Standard Q RADT performed reliably for patients with viral loads over 5.4 log_10_ or below Ct 27, which is in accordance to recent studies *(12, 14, 24)*. Since a virus concentration of 6 log_10_ copies/ml is commonly suspected to be the threshold for contagiousness of the patient, we aimed to investigate the correlation between RADT result and SARS-CoV-2 in vitro infectivity *(17, 18)*.

In contrast to highly sensitive nucleic acid-based detection methods that do not specifically test for intact viral particles required for transmission, viral culture is a frequently used, albeit laborious, method to determine the presence of infectious virus in clinical samples *(25–28)*. Although the detection of viable virus in cell culture models is strong evidence of infectiousness, a negative result does not eliminate the possibility of human transmission *(11, 29)*. Moreover, the validity of viral culture as a surrogate for infectivity may depend on the susceptibility of the cell line used *(26–28)*. However, loss of infectious titer in classical Vero E6 cells has been associated with a lack of transmission despite detection of viral RNA in preclinical models *(30)*.

To investigate whether the Standard Q RADT might be able to reliably detect culture-positive samples in Vero E6 cells, we attempted virus isolation from samples positive for SARS-CoV-2 RNA. All samples from which virus could be recovered had previously tested positive in RADT. Moreover, none of the samples tested negative by RADT contained infectious virus determined by cell culture. Furthermore, when taking symptom duration into account, we detected no positive culture after 6 days of symptom onset indicating a decreased probability of recovering viable virus as symptom duration increases *(18, 25, 31–33)*; at the same time the RADT identified positive samples for up to 9 days. While some groups have described virus isolation from samples above Ct 30 *(27, 34)*, here, positive cultures were only observed from samples with higher RNA loads which was in accordance with previous observations *(10, 24, 32, 35)*. Taking the time of suspected exposure and duration of symptoms into account *(27, 29)*, our results suggest that RADT testing is of potential use for determining infectivity at the time of sampling.

This study, however, is subject to some limitations. Although the examined single-center study population was large, our cohort might not be considered representative of the general population due to young age and disproportionate gender distribution. The data on symptoms and their duration are only reliable to a limited extent, since they were retrospectively analyzed from mostly self-reported symptoms entered into a web tool. Furthermore, instead of a nasal swab, we used an oro-and nasopharyngeal swab to investigate RADT performance, which impedes the feasibility for the general public.

In combating overdispersed SARS-CoV-2 transmission, rapid detection and isolation of highly infectious individuals is a primary goal *(8, 9, 36–40)*. In our investigation the Standard Q RADT was able to reliably detect high viral load as well as all culture positive samples. Therefore, this test could be used as a fast surrogate marker for viral cultivation in order to identify and prevent SARS-CoV-2 transmissions by highly infectious individuals. Although less sensitive than RT-qPCR, RADT could compensate for this disadvantage through easy and feasible mass screenings *(8, 41)*. Furthermore, one might suspect that RT-qPCR positive, but RADT negative individuals do not pose a high risk of transmissions, since all samples remained culture negative in our experimental setup. However, individual results must be interpreted with caution as SARS-CoV-2 infection could remain undetected in early stages. Simple to perform and applicable anywhere, RADT enable mass testing as a complementary method to RT-qPCR to more effectively combat the SARS-CoV-2 pandemic.

## MATERIALS AND METHODS

### Study design

Between 26^th^ October 2020 and 08^th^ January 2021 all enrolled participants were tested for SARS-CoV-2 infection by RT-qPCR at the University Hospital Cologne, Department I of Internal Medicine, Cologne, Germany. SARS-CoV-2 testing was available for individuals from the general population with COVID-19 symptoms, suspected disease or SARS-CoV-2 exposure as part of routine diagnostics, as well as for hospital staff members as part of screening measures. For quality control, an RADT was simultaneously performed after verbal consent. Specimens from two combined oro-and nasopharyngeal swabs were obtained by the same trained personnel and transferred into virus transport and preservation medium (biocomma®, Shenzhen, China) or BD ESwab™ (Becton & Dickinson, Sparks, MD, USA). All samples were routinely processed within 12 hours after collection, while the RADT was performed immediately on site using the second sample. Upon approval by the Institutional Review Board of the University of Cologne, results were retrospectively analyzed including clinical data retrieved from a symptoms diary webtool that all individuals registering for a SARS-CoV-2 test are asked to complete. Due to a number of implausible self-reported entries, only webtool entries not older than a week from the time of testing were included into symptom analysis. Patients were categorized as symptomatic, if reported symptom duration at the time of testing was ≤ 14 days and one of the following symptoms was found: fever, cough, rhinorrhea, nausea, diarrhea, shortness of breath, and/or a new olfactory or taste disorder.

### Rapid antigen detection test

The Standard Q COVID-19 Ag Test (SD Biosensor Inc., Suwon-si, Republic of Korea/Hoffmann La Roche AG, Basel, Switzerland) was performed according to the manufacturer’s instructions using the enclosed dry swab with one modification (Noble Biosciences, Inc., Hwaseong-si, Republic of Korea). Instead of a nasopharyngeal swab, a combined oro-and nasopharyngeal swab was performed to ensure comparability with RT-qPCR. Operating instructions in brief: The collected swab was mixed in the provided tube of collection medium and 3 drops were applied through a nozzle cap onto the test strip. Results were read out visually after 15-20 minutes by medically trained and instructed personnel. In accordance with the manufacturer’s reference guide, faint lines were considered positive if the control line was also present.

### Real time reverse transcription PCR

RT-qPCR was performed using different SARS-CoV-2 RNA detection protocols that were normalized according to the same standard. The following SARS-CoV-2 detection protocols were utilized: (1) Nucleic acid extraction was done for 935 (46.10%) samples using the MagNA Pure 96® system DNA and viral NA Large Volume Kit (Roche Diagnostics, Mannheim, Germany) according to the manufacturer’s instructions. After RNA purification from 500 µl viral transport medium and elution into 100 µl elution buffer, RT-qPCR was performed using the LightMix® SarbecoV E-gene plus equine arteritis virus (EAV) control kit (TIB Molbiol, Berlin, Germany) and an inhouse N-gene primer set in multiplex RT-qPCR adapted to the manufacturer’s instructions on a LightCycler® 480 II System (Roche Diagnostics, Mannheim, Germany). (2) cobas® SARS-CoV-2 Test kit running on the cobas 6800® (Roche Diagnostics, Mannheim, Germany) was used for 407 (20.07%) samples targeting the viral E-gen and ORF1a/b regions according to the manufacturer’s instructions. (3) SARS-CoV-2 AMP Kit running on the Alinity m (Abbott, Illinois, USA) was used for 63 (3.11%) specimens targeting the viral N-and RdRp-genes according to the manufacturer’s instructions. (4) Multiplex RT-qPCR using the LightMix® SarbecoV E-gene (TIB Molbiol, Berlin, Germany), an inhouse N-gene primer/probe set and a human β-globin primer set as internal control, running on the Panther Fusion® (Hologic, Wiesbaden, Germany) was used for 36 (1.78%) samples. (5) For 579 (28.55%) specimens, samples from up to ten asymptomatic employees were pooled and tested for SARS-CoV-2 using the methods described in (1), (2) and (3). Positive pools were resolved and samples were tested separately as described in (1) to (4). (6) Xpert® Xpress SARS-CoV-2 (Cepheid, Sunnyvale, USA) test kit was used for 8 (0.39%) samples according to the manufacturer’s instructions. To enable comparison of cycle threshold (Ct) values obtained by the different RT-qPCR methods, Ct values were translated into copies/ml and then converted to a cobas 6800® adjusted Ct value. For this purpose, a standard curve was extrapolated by a regression model using seven serial dilutions from a high titer SARS-CoV-2 sample that were tested with all five detection methods described above. For final adjustments to the model, two SARS-CoV-2 samples from INSTAND -Society for the Promotion of Quality Assurance in medical laboratories e.V. (Düsseldorf, Germany; in cooperation with the Robert Koch-Institute and the Institute of Virology, Charité, Berlin) were used for Ct-based absolute RNA quantification.

### SARS-CoV-2 Culture

Vero E6 cells (ATCC CRL-1586) were cultured in complete medium (CM) consisting of Dulbecco’s modified Eagle Medium (DMEM; Thermo Fisher Scientific-Gibco, Waltham, MA) supplemented with 10% fetal calf serum (FCS; GE Healthcare, Chicago, IL), 200 units/ml Penicillin, 200 µg/ml Streptomycin, 0.25 µg/ml Amphotericin B, 2 mM L-Glutamine and 1 mM sodium pyruvate (all by Thermo Fisher Scientific-Gibco, Waltham, MA) at 37°C in an incubator with 5% CO_2_. One day prior to infection 0.3 x 10e6 cells were seeded onto T25 flasks in 5 ml of CM. Retained swab samples were stored for up to 8 hours at 4°C in transport medium until RT-qPCR results became available. For virus cultures, 250 µl of SARS-CoV-2 RT-qPCR positive samples were diluted 1:5 in infection medium (IM) consisting of complete medium with FCS reduced to 2%. After removing the cell culture supernatant, the diluted samples were added to Vero E6 cells and incubated for one hour at 37°C and 5% CO_2_. After washing with 5 ml of PBS (Thermo Fisher Scientific-Gibco, Waltham, MA), 5 ml of IM was added and cells were cultured for 7 days at 37°C and 5% CO_2_. Cells were checked for presence of cytopathic effects (CPE) on day 4 and day 7. On both days 1 ml of culture supernatant was harvested and stored at −80°C for conformation of positive cultures through RT-qPCR as described in 2.3 (1). All virus isolation experiments were performed under BSL-3 conditions.

### Dilution Series

250 µl retained samples of patients with high RNA load identified by RT-qPCR were thawed and serially diluted in IM (1:5). Virus cultivation and RT-qPCR were performed as described in section 2.3 and 2.4, respectively. RADT were performed with 250 µl of original samples and each dilution as described above.

### Statistical analysis

Sensitivity and Specificity with 95% confidence intervals (CI) as well as positive and negative prediction values were calculated using the RT-qPCR as a reference. Culture and RADT results were evaluated by a contingency table and a p-value was calculated with Fisher’s exact test. Mann-Whitney U-test (MWU) was used to compare differences between medians. P values <0.05 were considered significant. Probit regression was carried out using a generalized linear model (R-function glm) with probit link function. To correct for repeated measurements from the same individual on different days, the basic analyses were modified as follows. (i) Confusion matrices for the calculation of all performance measures were calculated in a weighted manner so that the contribution of a single test is inversely proportional to the number of tests taken from the corresponding individual. (ii) For comparing two populations of data points, a generalized estimating equation (R-function gee::gee) was fitted using each patient as its own cluster and an exchangeable correlation structure. (iii) Probit regression was carried out by fitting a generalized linear mixed model with probit link function and random intercepts for each individual (R-function GLMMadaptive::mixed_model). Marginal means and confidence bands were calculated with R-function ggeffects::ggpredict. Data analysis was performed using Microsoft Excel 16.44 (Microsoft), GraphPad Prism 9 (GraphPad Software, Inc.), Python 3.8.3, and R 3.6.3.

## Data Availability

All data, code, and materials used in the analysis will be made available upon request.

## Ethics

The Institutional Review Board of the University of Cologne acknowledged and approved the study under application number 21-1039.

## Supplementary Materials

Table S1. Additional Cohort Description

Table S2. RADT results by symptoms and follow-up testing

Fig. S1. Virus cultivation of 1:5 dilutions.

Fig. S2. Modified data analyses to correct for repeated testing of single individuals.

## Acknowledgments

We thank all study participants who devoted time to our research; members of the Klein Laboratory and the University Hospital Cologne’s Institute of Virology and SARS-CoV-2 diagnostic center for continuous support and helpful discussions; Daniela Weiland and Nadine Henn for lab management and assistance; and the Becker Laboratory, Marburg, for sharing Vero E6 cells.

## Funding

Bundesministerium für Bildung und Forschung grant 01KX2021 („B-FAST”, „NaDoUniMedCovid19”)

German Center for Infection Research (MK)

Köln Fortune Program (NP, MM)

## Author contributions

Conceptualization: MK, NP, MM, FK

Methodology: MK, NP, MM, KV, HG, EH, FK

Investigation: MK, NP, MM, RE, MW, IF, FD, LG, CL, GF, HG

Visualization: MK, NP

Funding acquisition: NPfeifer, FK

Project administration: MK, NP

Supervision: EH, Npfeifer, FK

Writing – original draft: MK, NP

Writing – review & editing: MK, NP, MM, KV, RE, HG, FK

## Competing interests

The Rapid Antigen Detection Tests investigated in this study were kindly provided by Hoffmann-La Roche AG (Basel, Switzerland).

## Data and materials availability

All data, code and materials used in the analysis will be made available upon publication.

## Notes

### Funding Statement

Bundesministerium fuer Bildung und Forschung grant 01KX2021 (B-FAST, NaDoUniMedCovid19)                                                                                          
German Center for Infection Research (MK)
Koeln Fortune Program (NP, MM)

### Summary of Updates

Updated title; Clarified number of individuals with multiple testing in cohort description; Evaluated RADT performance using only the first swab of each individual; Added panel in Figure S2 and updated results part for repeated testing to include additional analysis

## References and Notes

1. C. Fraser, S. Riley, R. M. Anderson, N. M. Ferguson, Factors that make an infectious disease outbreak controllable, Proceedings of the National Academy of Sciences 101, 6146–6151 (2004).

2. J. Hellewell, S. Abbott, A. Gimma, N. I. Bosse, C. I. Jarvis, T. W. Russell, J. D. Munday, A. J. Kucharski, W. J. Edmunds, S. Funk, R. M. Eggo, F. Sun, S. Flasche, B. J. Quilty, N. Davies, Y. Liu, S. Clifford, P. Klepac, M. Jit, C. Diamond, H. Gibbs, K. van Zandvoort, Feasibility of controlling COVID-19 outbreaks by isolation of cases and contacts, The Lancet Global Health, S2214109X20300747 (2020).

3. M. E. Kretzschmar, G. Rozhnova, M. C. J. Bootsma, M. van Boven, J. H. H. M. van de Wijgert, M. J. M. Bonten, Impact of delays on effectiveness of contact tracing strategies for COVID-19: a modelling study, The Lancet Public Health 5, e452–e459 (2020).

4. V. M. Corman, O. Landt, M. Kaiser, R. Molenkamp, A. Meijer, D. K. Chu, T. Bleicker, S. Brünink, J. Schneider, M. L. Schmidt, D. G. Mulders, B. L. Haagmans, B. van der Veer, S. van den Brink, L. Wijsman, G. Goderski, J.-L. Romette, J. Ellis, M. Zambon, M. Peiris, H. Goossens, C. Reusken, M. P. Koopmans, C. Drosten, Detection of 2019 novel coronavirus (2019-nCoV) by real-time RT-PCR, Eurosurveillance 25 (2020), doi:10.2807/1560-7917.ES.2020.25.3.2000045.

5. J. Sun, J. Xiao, R. Sun, X. Tang, C. Liang, H. Lin, L. Zeng, J. Hu, R. Yuan, P. Zhou, J. Peng, Q. Xiong, F. Cui, Z. Liu, J. Lu, J. Tian, W. Ma, C. Ke, Prolonged Persistence of SARS-CoV-2 RNA in Body Fluids, Emerg. Infect. Dis. 26, 1834–1838 (2020).

6. M. Cevik, K. Kuppalli, J. Kindrachuk, M. Peiris, Virology, transmission, and pathogenesis of SARS-CoV-2, BMJ, m3862 (2020).

7. C.-G. Huang, K.-M. Lee, M.-J. Hsiao, S.-L. Yang, P.-N. Huang, Y.-N. Gong, T.-H. Hsieh, P.-W. Huang, Y.-J. Lin, Y.-C. Liu, K.-C. Tsao, S.-R. Shih, A. J. McAdam, Ed. Culture-Based Virus Isolation To Evaluate Potential Infectivity of Clinical Specimens Tested for COVID-19, J Clin Microbiol 58, e01068–20, /jcm/58/8/JCM.01068-20.atom (2020).

8. M. J. Mina, R. Parker, D. B. Larremore, Rethinking Covid-19 Test Sensitivity — A Strategy for Containment, N Engl J Med 383, e120 (2020).

9. M. J. Mina, T. E. Peto, M. García-Fiñana, M. G. Semple, I. E. Buchan, Clarifying the evidence on SARS-CoV-2 antigen rapid tests in public health responses to COVID-19, The Lancet, S0140673621004256 (2021).

10. I. W. Pray, L. Ford, D. Cole, C. Lee, J. P. Bigouette, G. R. Abedi, D. Bushman, M. J. Delahoy, D. Currie, B. Cherney, M. Kirby, G. Fajardo, M. Caudill, K. Langolf, J. Kahrs, P. Kelly, C. Pitts, A. Lim, N. Aulik, A. Tamin, J. L. Harcourt, K. Queen, J. Zhang, B. Whitaker, H. Browne, M. Medrzycki, P. Shewmaker, J. Folster, B. Bankamp, M. D. Bowen, N. J. Thornburg, K. Goffard, B. Limbago, A. Bateman, J. E. Tate, D. Gieryn, H. L. Kirking, R. Westergaard, M. Killerby, CDC COVID-19 Surge Laboratory Group, CDC COVID-19 Surge Laboratory Group, B. Jiang, J. Vinjé, A. L. Hopkins, E. Katz, L. Barclay, M. Esona, R. Gautam, S. Mijatovic-Rustempasic, S.-S. Moon, T. Bessey, P. Chhabra, S. L. Smart, R. Anderson, K. W. Radford, G. Kim, D. Thompson, C. Miao, M. Chen, L. Gade, R. Galloway, K. Sahibzada, N. M. Tran, S. Velusamy, H. Zheng, K. Nguyen, C. Hartloge, B. Jenkins, P. Wong, Performance of an Antigen-Based Test for Asymptomatic and Symptomatic SARS-CoV-2 Testing at Two University Campuses — Wisconsin, September–October 2020, MMWR Morb. Mortal. Wkly. Rep. 69, 1642– 1647 (2021).

11. J. L. Prince-Guerra, O. Almendares, L. D. Nolen, J. K. L. Gunn, A. P. Dale, S. A. Buono, M. Deutsch-Feldman, S. Suppiah, L. Hao, Y. Zeng, V. A. Stevens, K. Knipe, J. Pompey, C. Atherstone, D. P. Bui, T. Powell, A. Tamin, J. L. Harcourt, P. L. Shewmaker, M. Medrzycki, P. Wong, S. Jain, A. Tejada-Strop, S. Rogers, B. Emery, H. Wang, M. Petway, C. Bohannon, J. M. Folster, A. MacNeil, R. Salerno, W. Kuhnert-Tallman, J. E. Tate, N. J. Thornburg, H. L. Kirking, K. Sheiban, J. Kudrna, T. Cullen, K. K. Komatsu, J. M. Villanueva, D. A. Rose, J. C. Neatherlin, M. Anderson, P. A. Rota, M. A. Honein, W. A. Bower, Evaluation of Abbott BinaxNOW Rapid Antigen Test for SARS-CoV-2 Infection at Two Community-Based Testing Sites — Pima County, Arizona, November 3–17, 2020, MMWR Morb. Mortal. Wkly. Rep. 70, 100–105 (2021).

12. N. Kohmer, T. Toptan, C. Pallas, O. Karaca, A. Pfeiffer, S. Westhaus, M. Widera, A. Berger,. Hoehl, M. Kammel, S. Ciesek, H. F. Rabenau, The Comparative Clinical Performance of Four SARS-CoV-2 Rapid Antigen Tests and Their Correlation to Infectivity In Vitro, JCM 10, 328 (2021).

13. A. Nalumansi, T. Lutalo, J. Kayiwa, C. Watera, S. Balinandi, J. Kiconco, J. Nakaseegu, D. Olara, E. Odwilo, J. Serwanga, B. Kikaire, D. Ssemwanga, S. Nabadda, I. Ssewanyana, D. Atwine, H. Mwebesa, H. K. Bosa, C. Nsereko, M. Cotten, R. Downing, J. Lutwama, P. Kaleebu, Field Evaluation of the Performance of a SARS-CoV-2 Antigen Rapid Diagnostic Test in Uganda using Nasopharyngeal Samples, International Journal of Infectious Diseases, S120197122032275X (2020).

14. G. Pilarowski, C. Marquez, L. Rubio, J. Peng, J. Martinez, D. Black, G. Chamie, D. Jones, J. Jacobo, V. Tulier-Laiwa, S. Rojas, S. Rojas, C. Cox, M. Petersen, J. DeRisi, D. V. Havlir, Field performance and public health response using the BinaxNOW TM Rapid SARS-CoV-2 antigen detection assay during community-based testing, Clinical Infectious Diseases, ciaa1890 (2020).

15. E. Albert, I. Torres, F. Bueno, D. Huntley, E. Molla, M.Á. Fernández-Fuentes, M. Martínez, S. Poujois, L. Forqué, A. Valdivia, C. Solano de la Asunción, J. Ferrer, J. Colomina, D. Navarro, Field evaluation of a rapid antigen test (Panbio™ COVID-19 Ag Rapid Test Device) for COVID-19 diagnosis in primary healthcare centres, Clinical Microbiology and Infection 27, 472.e7-472.e10 (2021).

16. M. Marks, P. Millat-Martinez, D. Ouchi, C. h Roberts, A. Alemany, M. Corbacho-Monné, M. Ubals, A. Tobias, C. Tebé, E. Ballana, Q. Bassat, B. Baro, M. Vall-Mayans, C. G-Beiras, N. Prat, J. Ara, B. Clotet, O. Mitjà, Transmission of COVID-19 in 282 clusters in Catalonia, Spain: a cohort study, The Lancet Infectious Diseases, S1473309920309853 (2021).

17. J. J. A. van Kampen, D. A. M. C. van de Vijver, P. L. A. Fraaij, B. L. Haagmans, M. M. Lamers, N. Okba, J. P. C. van den Akker, H. Endeman, D. A. M. P. J. Gommers, J. J. Cornelissen, R. A. S. Hoek, M. M. van der Eerden, D. A. Hesselink, H. J. Metselaar, A. Verbon, J. E. M. de Steenwinkel, G. I. Aron, E. C. M. van Gorp, S. van Boheemen, J. C. Voermans, C. A. B. Boucher, R. Molenkamp, M. P. G. Koopmans, C. Geurtsvankessel, A. A. van der Eijk, Duration and key determinants of infectious virus shedding in hospitalized patients with coronavirus disease-2019 (COVID-19), Nat Commun 12, 267 (2021).

18. R. Wölfel, V. M. Corman, W. Guggemos, M. Seilmaier, S. Zange, M. A. Müller, D. Niemeyer, T. C. Jones, P. Vollmar, C. Rothe, M. Hoelscher, T. Bleicker, S. Brünink, J. Schneider, R. Ehmann, K. Zwirglmaier, C. Drosten, C. Wendtner, Virological assessment of hospitalized patients with COVID-2019, Nature 581, 465–469 (2020).

19. B. Diao, K. Wen, J. Zhang, J. Chen, C. Han, Y. Chen, S. Wang, G. Deng, H. Zhou, Y. Wu, Accuracy of a nucleocapsid protein antigen rapid test in the diagnosis of SARS-CoV-2 infection, Clinical Microbiology and Infection 27, 289.e1-289.e4 (2021).

20. M. Linares, R. Pérez-Tanoira, A. Carrero, J. Romanyk, F. Pérez-García, P. Gómez-Herruz, T. Arroyo, J. Cuadros, Panbio antigen rapid test is reliable to diagnose SARS-CoV-2 infection in the first 7 days after the onset of symptoms, Journal of Clinical Virology 133, 104659 (2020).

21. S. Lambert-Niclot, A. Cuffel, S. Le Pape, C. Vauloup-Fellous, L. Morand-Joubert, A.-M. Roque-Afonso, J. Le Goff, C. Delaugerre, A. J. McAdam, Ed. Evaluation of a Rapid Diagnostic Assay for Detection of SARS-CoV-2 Antigen in Nasopharyngeal Swabs, J Clin Microbiol 58, e00977-20, /jcm/58/8/JCM.00977-20.atom (2020).

22. F. Cerutti, E. Burdino, M. G. Milia, T. Allice, G. Gregori, B. Bruzzone, V. Ghisetti, Urgent need of rapid tests for SARS CoV-2 antigen detection: Evaluation of the SD-Biosensor antigen test for SARS-CoV-2, Journal of Clinical Virology 132, 104654 (2020).

23. G. Turcato, A. Zaboli, N. Pfeifer, L. Ciccariello, S. Sibilio, G. Tezza, D. Ausserhofer, Clinical application of a rapid antigen test for the detection of SARS-CoV-2 infection in symptomatic and asymptomatic patients evaluated in the emergency department: a preliminary report., Journal of Infection, S0163445320307738 (2020).

24. A. Strömer, R. Rose, M. Schäfer, F. Schön, A. Vollersen, T. Lorentz, H. Fickenscher, A. Krumbholz, Performance of a Point-of-Care Test for the Rapid Detection of SARS-CoV-2 Antigen, Microorganisms 9, 58 (2020).

25. Guidance for discharge and ending of isolation of people with COVID-19 (European Centre for Disease Prevention and Control, Stockholm, 2020; https://www.ecdc.europa.eu/sites/default/files/documents/Guidance-for-discharge-and-ending-of-isolation-of-people-with-COVID-19.pdf).

26. A. Pekosz, V. Parvu, M. Li, J. C. Andrews, Y. C. Manabe, S. Kodsi, D. S. Gary, C. Roger-Dalbert, J. Leitch, C. K. Cooper, Antigen-Based Testing but Not Real-Time Polymerase Chain Reaction Correlates With Severe Acute Respiratory Syndrome Coronavirus 2 Viral Culture, Clinical Infectious Diseases, ciaa1706 (2021).

27. T. Jefferson, E. A. Spencer, J. Brassey, C. Heneghan, Viral cultures for COVID-19 infectious potential assessment – a systematic review, Clinical Infectious Diseases, ciaa1764 (2020).

28. K. Basile, K. McPhie, I. Carter, S. Alderson, H. Rahman, L. Donovan, S. Kumar, T. Tran, D. Ko, T. Sivaruban, C. Ngo, C. Toi, M. V. O’Sullivan, V. Sintchenko, S. C.-A. Chen, S. Maddocks, D. E. Dwyer, J. Kok, Cell-based culture of SARS-CoV-2 informs infectivity and safe de-isolation assessments during COVID-19, Clinical Infectious Diseases, ciaa1579 (2020).

29. C. Rhee, S. Kanjilal, M. Baker, M. Klompas, Duration of Severe Acute Respiratory Syndrome Coronavirus 2 (SARS-CoV-2) Infectivity: When Is It Safe to Discontinue Isolation?, Clinical Infectious Diseases, ciaa1249 (2020).

30. S. F. Sia, L.-M. Yan, A. W. H. Chin, K. Fung, K.-T. Choy, A. Y. L. Wong, P. Kaewpreedee, R. A. P. M. Perera, L. L. M. Poon, J. M. Nicholls, M. Peiris, H.-L. Yen, Pathogenesis and transmission of SARS-CoV-2 in golden hamsters, Nature 583, 834–838 (2020).

31. M. Cevik, M. Tate, O. Lloyd, A. E. Maraolo, J. Schafers, A. Ho, SARS-CoV-2, SARS-CoV, and MERS-CoV viral load dynamics, duration of viral shedding, and infectiousness: a systematic review and meta-analysis, The Lancet Microbe 2, e13–e22 (2021).

32. J. Bullard, K. Dust, D. Funk, J. E. Strong, D. Alexander, L. Garnett, C. Boodman, A. Bello, A. Hedley, Z. Schiffman, K. Doan, N. Bastien, Y. Li, P. G. Van Caeseele, G. Poliquin, Predicting infectious SARS-CoV-2 from diagnostic samples, Clinical Infectious Diseases, ciaa638 (2020).

33. R. A. P. M. Perera, E. Tso, O. T. Y. Tsang, D. N. C. Tsang, K. Fung, Y. W. Y. Leung, A. W. H. Chin, D. K. W. Chu, S. M. S. Cheng, L. L. M. Poon, V. W. M. Chuang, M. Peiris, SARS-CoV-2 Virus Culture and Subgenomic RNA for Respiratory Specimens from Patients with Mild Coronavirus Disease, Emerg. Infect. Dis. 26, 2701–2704 (2020).

34. V. Gniazdowski, C. P. Morris, S. Wohl, T. Mehoke, S. Ramakrishnan, P. Thielen, H. Powell, B. Smith, D. T. Armstrong, M. Herrera, C. Reifsnyder, M. Sevdali, K. C. Carroll, A. Pekosz, H.H. Mostafa, Repeat COVID-19 Molecular Testing: Correlation of SARS-CoV-2 Culture with Molecular Assays and Cycle Thresholds, Clinical Infectious Diseases, ciaa1616 (2020).

35. M.-C. Kim, C. Cui, K.-R. Shin, J.-Y. Bae, O.-J. Kweon, M.-K. Lee, S.-H. Choi, S.-Y. Jung, M.-S. Park, J.-W. Chung, Duration of Culturable SARS-CoV-2 in Hospitalized Patients with Covid-19, N Engl J Med, NEJMc2027040 (2021).

36. D. B. Larremore, B. Wilder, E. Lester, S. Shehata, J. M. Burke, J. A. Hay, M. Tambe, M. J. Mina, R. Parker, Test sensitivity is secondary to frequency and turnaround time for COVID-19 screening, Sci. Adv. 7, eabd5393 (2021).

37. N. Muller, M. Kunze, F. Steitz, N. J. Saad, B. Mühlemann, J. I. Beheim-Schwarzbach, J. Schneider, C. Drosten, L. Murajda, S. Kochs, C. Ruscher, J. Walter, N. Zeitlmann, V. M. Corman, Severe Acute Respiratory Syndrome Coronavirus 2 Outbreak Related to a Nightclub, Germany, 2020, Emerg. Infect. Dis. 27, 645–648 (2020).

38. C.L. Correa-Martínez, S. Kampmeier, P. Kümpers, V. Schwierzeck, M. Hennies, W. Hafezi, J. Kühn, H. Pavenstädt, S. Ludwig, A. Mellmann, A. J. McAdam, Ed. A Pandemic in Times of Global Tourism: Superspreading and Exportation of COVID-19 Cases from a Ski Area in Austria, J Clin Microbiol 58, e00588-20, /jcm/58/6/JCM.00588-20.atom (2020).

39. D. C. Adam, P. Wu, J. Y. Wong, E. H. Y. Lau, T. K. Tsang, S. Cauchemez, G. M. Leung, B.J. Cowling, Clustering and superspreading potential of SARS-CoV-2 infections in Hong Kong, Nat Med 26, 1714–1719 (2020).

40. M. Cevik, J. L. Marcus, C. Buckee, T. C. Smith, Severe Acute Respiratory Syndrome Coronavirus 2 (SARS-CoV-2) Transmission Dynamics Should Inform Policy, Clinical Infectious Diseases, ciaa1442 (2020).

41. J. O. Lloyd-Smith, S. J. Schreiber, P. E. Kopp, W. M. Getz, Superspreading and the effect of individual variation on disease emergence, Nature 438, 355–359 (2005).

