## Supplementary Materials for "Assessment of SARS-CoV-2 infectivity by a Rapid Antigen Detection Test"

| | | Total | $PCR^+$ | $PCR^-$ | p-value |
|---|---|---|---|---|---|
| N | | 2,028 | 210 (10.26%) | 1,818 (89.74%) | - |
| Median Age (IQR) | | 32.25 (26.14-43.12) | 31.54 (24.40 - 46.47) | 32.31 (26.30 - 42.90) | - |
| Sex | m | 789 (38.90%) | 89 (42.38%) | 700 (38.50%) | 0.2954 (0.1166) |
| | w | 1,239 (61.10%) | 121 (57.62%) | 1,118 (61.50%) | |
| N | | 1,676 | 210 | 1,466 | - |
| Symptomatic | yes | 866 (51.67%) | 130 (61.90%) | 736 (50.20%) | 0.0015 (0.0013) |
| | no | 810 (48.33%) | 80 (38.10%) | 730 (49.80%) | |
| | | **Total** | **$RADT^+$** | **$RADT^-$** | **p-value** |
| N | | 2,028 | 92 (4.54%) | 1936 (95.46%) | - |
| Median Age (IQR) | | 32.25 (26.14-43.12) | 31.71 (24.33-49.79) | 32.26 (26.17-42.87) | - |
| Sex | m | 789 (38.90%) | 43 (46.74%) | 746 (38.53%) | 0.1257 (0.0945) |
| | w | 1,239 (61.10%) | 49 (53.26%) | 1190 (61.47%) | |
| N | | 1676 | 92 | 1584 | - |
| Symptomatic | yes | 866 (51.67%) | 73 (79.34%) | 793 (50.06%) | <0.0001 (<0.0001) |
| | no | 810 (48.33%) | 19 (20.65%) | 791 (49.94%) | |
| | | **Total** | **$culture^+$** | **$culture^-$** | **p-value** |
| N | | 118 | 29 (24.58%) | 89 (75.42%) | - |
| Median Age (IQR) | | 32.12 (25.61-46.47) | 40.65 (30.01-52.15) | 31.38 (25.52-44.84) | - |
| Sex | m | 46 (38.98%) | 15 (51.72%) | 31 (34.83%) | 0.1270 (0.1133) |
| | w | 72 (61.02%) | 14 (48.28%) | 58 (65.17%) | |
| N | | 118 | 29 | 89 | - |
| Symptomatic | yes | 72 (61.02%) | 20 (68.97%) | 52 (58.43%) | 0.3833 (0.8183) |
| | no | 46 (38.98%) | 9 (31.03%) | 37 (41.57%) | |

**Table S1. Additional cohort description.** Test results by age, sex and symptoms (weighted analyses for repeated measures in brackets; all p-values by Fisher's exact test).

| | | Total | RADT+ | RADT- | p-value |
|---|---|---|---|---|---|
| N | | 1676 | 92 | 1584 | - |
| Symptomatic | yes | 866 | 73 | 793 | <0.0001 |
| | no | 810 | 19 | 791 | (<0.0001) |
| Cough | yes | 347 (41.02%) | 37 (58.73%) | 310 (39.59%) | 0.0034 |
| | no | 499 (58.98%) | 26 (41.27%) | 473 (60.41%) | (0.0059) |
| Fever | yes | 101 (11.94%) | 24 (38.10%) | 77 (9.83%) | <0.0001 |
| | no | 745 (88.06) | 39 (61.90%) | 706 (90.17%) | (<0.0001) |
| Rhinitis | yes | 466 (55.08%) | 41 (65.08%) | 425 (54.28%) | 0.1141 |
| | no | 380 (44.92%) | 22 (34.93%) | 358 (45.72%) | (0.1806) |
| Loss of taste/smell | yes | 83 (9.81%) | 35 (31.82%) | 48 (6.52%) | <0.0001 |
| | no | 763 (90.19%) | 75 (68.18%) | 688 (93.48%) | (<0.0001) |
| Headache | yes | 503 (59.46%) | 49 (77.78%) | 454 (57.98%) | 0.0020 |
| | no | 343 (40.54%) | 14 (22.22%) | 329 (42.02%) | (0.0024) |
| Sore throat | yes | 512 (60.52%) | 35 (55.56%) | 477 (60.92%) | 0.4232 |
| | no | 334 (39.48%) | 28 (44.44%) | 306 (39.08%) | (0.3407) |
| Limb pain | yes | 205 (24.23%) | 28 (44.44%) | 177 (22.61%) | 0.0003 |
| | no | 641 (75.77%) | 35 (55.56%) | 606 (77.39%) | (0.0003) |
| Respiratory problems | yes | 48 (5.67%) | 4 (6.35%) | 44 (5.62%) | 0.7757 |
| | no | 798 (94.33%) | 59 (93.65%) | 739 (94.38%) | (0.7736) |
| Diarrhea | yes | 97 (11.47%) | 9 (14.29%) | 88 (11.24%) | 0.4167 |
| | no | 749 (88.53%) | 54 (85.71%) | 695 (88.76%) | (0.6733) |
| N | | 860 | 70 | 790 | - |
| Days since symptom onset | 0-2 d | 494 (57.44%) | 45 (64.29%) | 449 (56.84%) | |
| | 3-7 d | 299 (34.77%) | 23 (32.86%) | 276 (34.94%) | 0.2213 (0.2597) |
| | 8-14 d | 67 (7.79%) | 2 (2.86%) | 65 (8.23%) | |
| prior test result (within 8 weeks before study) | yes | 666 (32.84%) | 17 (18.48%) | 649 (33.52%) | 0.0021 |
| | unknown | 1362 (67.16%) | 75 (81.52%) | 1287 (66.48%) | (0.0060) |

**Table S2. RADT results by symptoms and follow-up testing** (weighted analyses for repeated measures in brackets; all p-values by Fisher's exact test).

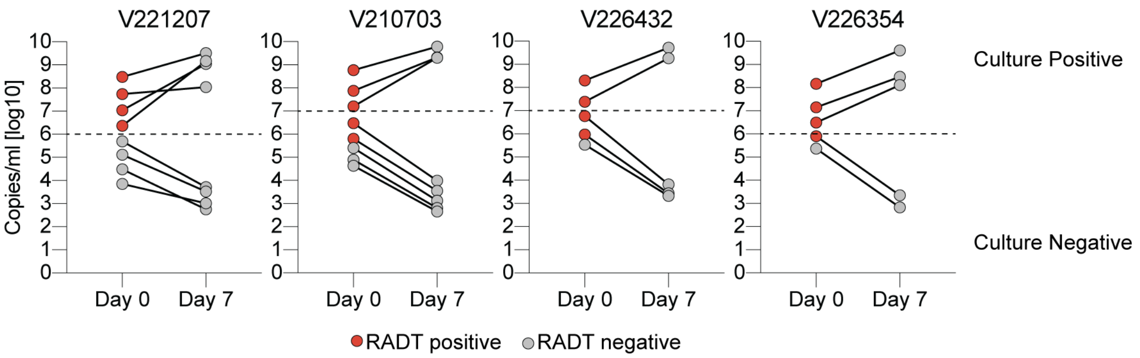


**Fig. S1. Virus cultivation of 1:5 dilutions.**
Samples of four different individuals with high viral loads were diluted and subsequently used for RADT testing and virus cultivation. Culture supernatant was tested in RT-qPCR on the day of inoculation and day 7 for detection of virus replication.

**A**

| | Original | Weighted | Simplified |
|---|---|---|---|
| Samples | 2028 | 1849 | 1849 |
| True negative | 1816 | 1657.67 | 1657 |
| True positive | 90 | 86.83 | 89 |
| False negative | 120 | 102.5 | 101 |
| False positive | 2 | 2 | 2 |
| Sensitivity | 0.43 | 0.46 | 0.47 |
| Specificity | 0.99 | 0.99 | 0.99 |

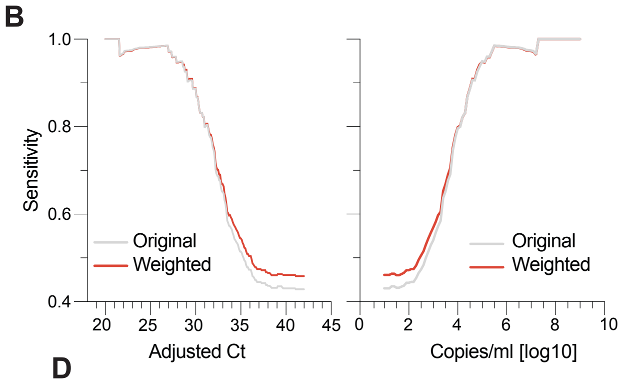


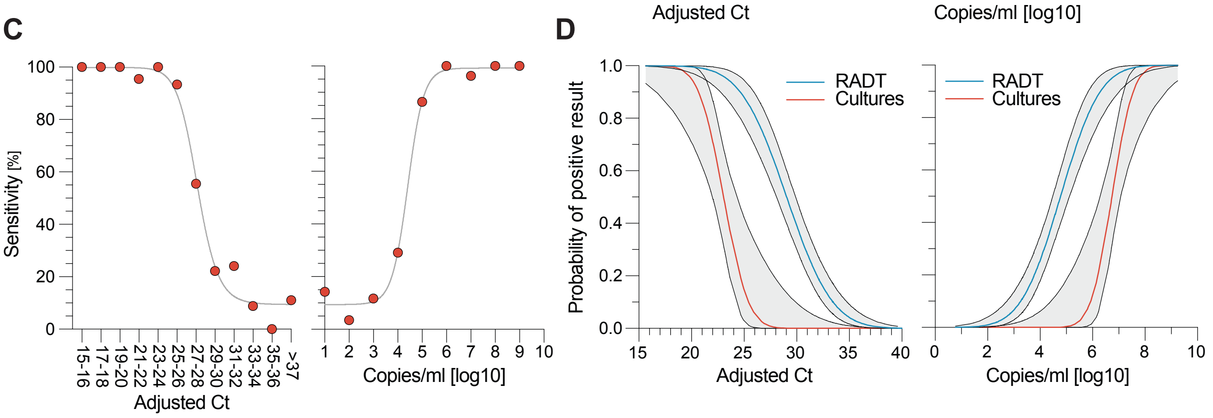


**Fig. S2. Modified data analyses to correct for repeated testing of single individuals.**
**(A)** Comparison of RADT performance data in either a weighted or simplified (only first swab result) fashion. **(B)** The cumulative sensitivity of the RADT is stratified by adjusted Ct values and RNA load in log copies/ml. **(C)** Sensitivity of RADT is stratified by adjusted Ct values and RNA load in log copies/ml for simplified analysis (only first swab result). **(D)** Probability of positive result for RADT and viral cultures in the context of adjusted Ct values and RNA load (Probit-Model, R-function GLMMadaptive). All weighted p-values remain $p < 0.0001$, Generalized Estimating Equations.
